# A multi-omics bidirectional mendelian randomization study and meta-analysis on the causal relationship between gut microbiota, inflammatory proteins, and fibromyalgia

**DOI:** 10.1101/2024.09.13.24313599

**Authors:** Mengqi Niu, Jing Li, Victoria Sarafian, Michael Maes

## Abstract

**Background:** Fibromyalgia (FM) is a chronic disorder characterized by widespread pain and immune dysregulation. Emerging evidence suggests that gut microbiota and inflammatory proteins may contribute to the development of FM.

**Objective:** The aim of this study was to investigate the causal relationships between gut microbiota, inflammatory proteins (cytokines/chemokines), and FM using bidirectional Mendelian randomization (MR) and meta-analysis approaches.

**Methods:** MR analyses were conducted using genetic data from European populations, employing methods such as MR-IVW, MR-Egger, and MR-weighted median. Reverse MR was also performed, with FM treated as the exposure. A meta-analysis was conducted to consolidate the findings.

**Results:** *Ruminococcus gauvreauii* was identified as a risk factor for FM, while *Enterorhabdus*, *Parabacteroides*, *Butyricicoccus*, and *Prevotella 9* were found to be protective. Five inflammatory proteins—C-X-C motif chemokine 5 (CXCL5), S100-A12, Leukemia inhibitory factor receptor (LIFR), Monocyte chemoattractant protein 2 (MCP-2/CCL8), and Tumor necrosis factor (TNF-α)—exhibited protective associations, while Natural killer cell receptor 2B4 (NKCR-2B4/CD244) and Interleukin-12 subunit beta (IL-12β) were associated with an increased risk of FM.

**Conclusion:** This study highlights the role of gut microbiota and inflammatory proteins (cytokines/chemokines) in the pathogenesis of FM. Through Gene Ontology (GO) functional enrichment and Kyoto Encyclopedia of Genes and Genomes (KEGG) pathway analyses, the findings suggest their involvement in immune regulation, inflammatory responses, and viral pathways. These findings provide new insights into potential therapeutic targets for modulating gut health and immune responses, opening new avenues for future research and clinical interventions.

## 1. Introduction

Fibromyalgia (FM) is a condition characterized by widespread chronic musculoskeletal pain, stiffness in specific areas, and a heightened sensitivity to pressure, known as tender points in soft tissue (1). Despite pain being its primary and prominent feature, FM is characterized by a complex array of symptoms including fatigue, sleep disturbances, and functional symptoms (medical symptoms that cannot be explained by structural or pathological definitions) (2). Globally, FM is estimated to affect 2.7% of the population, with higher prevalence rates observed in women, individuals over 50 years old, those with lower socioeconomic status, and obese individuals (3). However, due to the lack of consensus on the pathogenic mechanisms underpinning FM, there is currently no universally effective treatment (2).

A recent meta-analysis shows that FM is characterized by indicants of peripheral immune activation with changes in immunoregulatory cytokines and pro-inflammatory cytokines (4). FM patients exhibit enhanced circulating inflammatory cytokines and chemokines released by peripheral blood mononuclear cells (PBMCs), indicating activation of both innate and adaptive immunity (5–7). Current research revealed elevated concentrations of M1 macrophage cytokines, such as IL-6, IL-8, IL-1β, and TNF-α, in the serum of FM patients (4, 5, 8–10). Immune cells, including mast cells, monocytes, and neutrophils, which mediate the inflammatory process, may also contribute to the immune pathogenesis of FM (11). In patients with major depression, acute COVID-19 infection, Long-COVID, schizophrenia and end-stage renal disease, FM symptoms are associated with immune activation as indicated by increased levels of pro-inflammatory mediators (12–16). Interestingly, plasma levels of soluble gp130 (sgp130), the natural inhibitor of IL-6 transsignalling, is significantly decreased in FM (17), which together with increased IL-6 levels suggest that the immune activation in FM is mainly driven by increased IL-6 signaling (4).

Additionally, in animal studies, resident macrophages in muscle tissue have been shown to play a role in the development of chronic widespread muscle pain (18). Increasing evidence suggests that neurogenic inflammatory processes occurring in peripheral tissues, the spinal cord, and the brain contribute to the pathophysiology of FM (5, 19, 20). All of these translate into the clinical symptoms reported by FM patients, such as swelling and sensory dullness, which also impact central symptoms including cognitive changes and fatigue. Furthermore, physiological mechanisms related to stress and emotions are considered upstream driving factors of neurogenic inflammation in FM (7, 21).

In 2007, it was published that FM symptoms of chronic fatigue syndrome are associated with increased translocation of Gram-negative gut commensal bacteria, indicating leaky gut (22). In schizophrenia patients, FM symptoms are strongly associated with breakdown of the paracellular pathways (leaky gut) and increased bacterial translocation (13). Other findings suggested that gut dysbiosis may play a role in FM. Thus, studies by Minerbi et al. (23) and Minerbi and Fitzcharles et al. (24) reported associations between gut microbiota and FM. In addition, a systematic review showed that intestinal microbiota is involved in the pathophysiology of FM (25).

However, until now it has remained unclear whether there are causal relationships between immune activation, the gut-immune axis, and FM. Recently developed Mendelian randomization (MR) designs have been used to study whether specific exposures have causal effects on specific outcomes (26). As an analytical method, MR designs utilize lineage genetic variations (single nucleotide polymorphisms, SNPs) as instrumental variables for the target exposure (27). Based on the random assortment during meiosis and the fixed allocation of genetic variations at conception, MR studies are less susceptible to confounding factors and reverse causation biases. Additionally, the use of summary statistics from genome-wide association studies (GWAS) in two-sample MR designs significantly enhances the statistical power of causal inference (28). Therefore, in this study, we use summary statistics from GWAS to conduct MR research. The a priori hypothesis is to discover clear multi-omics causal relationships between FM and immune activation as assessed with cytokines/chemokines and changes in the gut microbiota indicating an inflammatory milieu or leaky gut.

## 2. Materials and Methods

### 2.1. Study Design

In our analysis, we used publicly available data from shared repositories, eliminating the need for additional ethical approval for this study. The MR analysis must meet three assumptions: (1) SNPs are associated with exposure; (2) SNPs are independent of confounders in the exposure-outcome relationship (independence assumption); (3) SNPs influence the outcome solely through exposure (29). **Figure 1** illustrates the workflow of this study.

**Figure 1.**
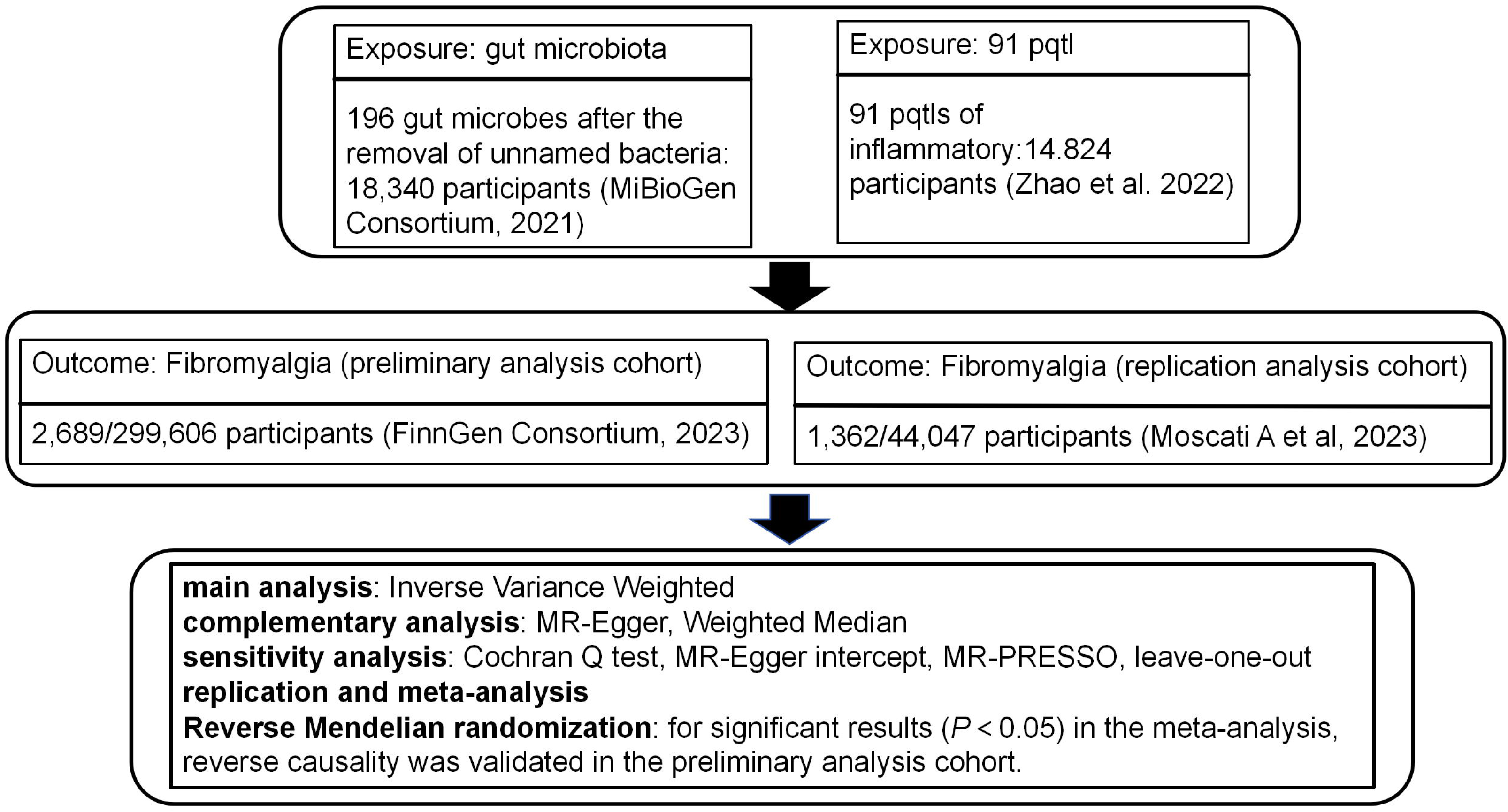
The flowchart of bidirectional mendelian randomization study and meta-analysis explore the causal relationship between gut microbiota, inflammatory proteins, and fibromyalgia. PQTL: protein quantitative trait loci

### 2.2. Data Sources

This study utilized three types of exposure data and two types of outcome data. To minimize bias due to population heterogeneity, only aggregated data from European populations were used. SNPs related to the human gut microbiome composition were obtained from the latest data from the MiBioGen international consortium (https://mibiogen.gcc.rug.nl/). This GWAS included 211 microbial taxa classified at six levels, from phylum to species (30). We sourced data on 91 inflammatory protein types from the study by Zhao et al (31). Summary statistics for FM outcomes were sourced from the FinnGen consortium (released in 2023, https://r10.finngen.fi/) and the study by Moscati et al (32).

### 2.3. Genetic Instrument Selection

(1) Based on previous studies, a significance threshold (p < 1×10[[) was applied for gut microbiota (33), while a stricter p-value (p < 5×10[[) was used for inflammatory proteins (34) to ensure a moderate number of SNPs for all traits. (2) To ensure independence of the instruments used for exposure, we excluded SNPs in linkage disequilibrium (LD) (r² < 0.001, clumping window = 10,000 kb) (35). (3) Palindromic SNPs with intermediate minor allele frequencies (MAF) were excluded (36). (4) To determine the existence of weak instrument bias, an F-statistic was computed to measure the power of the instrumental variable. An F-statistic above 10 indicated a low likelihood of weak instrument bias (37).

### 2.4. Two-Sample Mendelian Randomization

This study primarily relied on random effects inverse variance weighting (IVW) to assess the causal effect of gut microbiota, and inflammatory proteins on FM. The IVW estimates were derived by aggregating the Wald ratios of all genetic variants. IVW is based on the assumption that all SNPs do not exhibit horizontal pleiotropy, thus providing the most accurate causal effect estimate. To ensure robustness, we also employed MR-Egger and weighted median (WM) methods for supplementary analysis (28, 38).

Sensitivity analyses are essential for examining horizontal pleiotropy and heterogeneity, which can significantly impact MR estimates. In this study, we used three methods to detect and correct these issues: Cochran’s Q test, MR-Egger intercept test, and MR-PRESSO (Pleiotropy RESidual Sum and Outlier) analysis. A p-value < 0.05 from Cochran’s Q test indicated heterogeneity in the results (39). The MR-Egger intercept was used to detect directional pleiotropy and bias due to invalid instruments (38). Finally, MR-PRESSO was used to identify and correct for potential pleiotropic outliers (40). To assess the robustness of the results, we employed the leave-one-out method, sequentially excluding each SNP to determine if any single SNP drove the estimates (38).

We stringently screened the gut microbiota and cytokines/chemokines with potential causal relationships to FM based on several criteria: (1) Significant p-value (p < 0.05) from the primary MR IVW analysis; (2) Consistent direction and magnitude across three MR methods; (3) No significant heterogeneity or horizontal pleiotropy in MR results.

### 2.5. Meta-analysis and genetic directionality assessment

To comprehensively estimate the robustness of gut microbiota and inflammatory proteins identified by the aforementioned criteria, we used the FinnGen consortium’s summary statistics for FM as the preliminary analysis. We replicated the IVW analysis in another FM cohort (study by Moscati et al.). The meta-analysis of the results from the two MR analyses identified gut microbiota and immune proteins causally related to FM.

Additionally, to rule out bias from reverse causality and further evaluate the robustness of the causal relationship between gut microbiota and immune proteins, and FM, we conducted reverse MR analysis within the preliminary analysis cohort (FinnGen consortium) for significant results (P < 0.05) from the meta-analysis. This provided final evidence for causal relationships.

Statistical analyses were performed using R software (version 4.3.2) with the packages MendelianRandomization, MRPRESSO, TwoSampleMR, Meta, among others. For binary outcomes, results were reported as odds ratios (OR) with corresponding 95% confidence intervals (CI) to quantify the strength of causal relationships. For continuous outcomes, results were reported as β coefficients, standard errors (SE), and corresponding 95% CI.

### 2.6. Protein-protein interaction (PPI) annotation and enrichment

To better explore the interactions between proteins and the pathogenesis, we performed PPI annotation and enrichment analysis using STRING (https://cn.string-db.org) and Kyoto Encyclopedia of Genes and Genomes (KEGG) and Gene Ontology (GO) databases.

## 3. Results

### 3.1. Causal relationship between gut microbiota and fibromyalgia

After excluding unnamed bacteria, we identified 196 gut microbiotas. In the preliminary analysis cohort, six gut microbiota showed statistically significant results (**Table S1**), including family *Enterobacteriaceae* (OR = 0.68, 95% CI: 0.47-0.98, p = 0.041), genus *Butyricicoccus* (OR = 0.68, 95% CI: 0.49-0.95, p = 0.023), genus *Coprococcus1* (OR = 0.69, 95% CI: 0.52-0.93, p = 0.014), genus *Eggerthella* (OR = 1.31, 95% CI: 1.08-1.59, p = 0.006), genus *Ruminococcaceae* UCG005 (OR = 1.37, 95% CI: 1.04-1.81, p = 0.025), and order Enterobacteriales (OR = 0.68, 95% CI: 0.47-0.98, p = 0.041). Significant gut microbiota also passed sensitivity tests including Cochran’s Q test (p > 0.05), MR-Egger intercept test (p > 0.05), MR-PRESSO analysis (p > 0.05), and leave-one-out analysis (**Table S2, Figure S1**). We replicated the MR analysis using another GWAS dataset for FM. As expected, similar trends were observed for the candidate gut microbiota in another GWAS dataset (study by Moscati et al.), but results were not significant due to large sample size differences.

To strengthen the credibility of the estimates, we performed a meta-analysis of MR results from the preliminary and replication cohorts, identifying six gut microbiota influencing FM (**Figure 2**). Specifically, higher levels of genus *Enterorhabdus* (OR = 0.78, 95% CI: 0.61-1.00, p = 0.048), genus *Parabacteroides* (OR = 0.61, 95% CI: 0.41-0.91, p = 0.016), genus *Butyricicoccus* (OR = 0.73, 95% CI: 0.55-0.96, p = 0.027), genus *Candidatus Soleaferrea* (OR = 0.83, 95% CI: 0.70-1.00, p = 0.046), and genus *Prevotella 9* (OR = 0.83, 95% CI: 0.69-0.99, p = 0.035) reduced the risk of FM. However, higher levels of genus *Ruminococcus gauvreauii* group (OR = 1.56, 95% CI: 1.19-2.04, p = 0.001) increased susceptibility to FM. In the reverse MR analysis (**Table 1**), we found a reverse causal relationship between FM and the genus *Candidatus Soleaferrea* (p < 0.05), indicating that genus *Candidatus Soleaferrea* did not pass the reverse causality validation.

**Figure 2.**
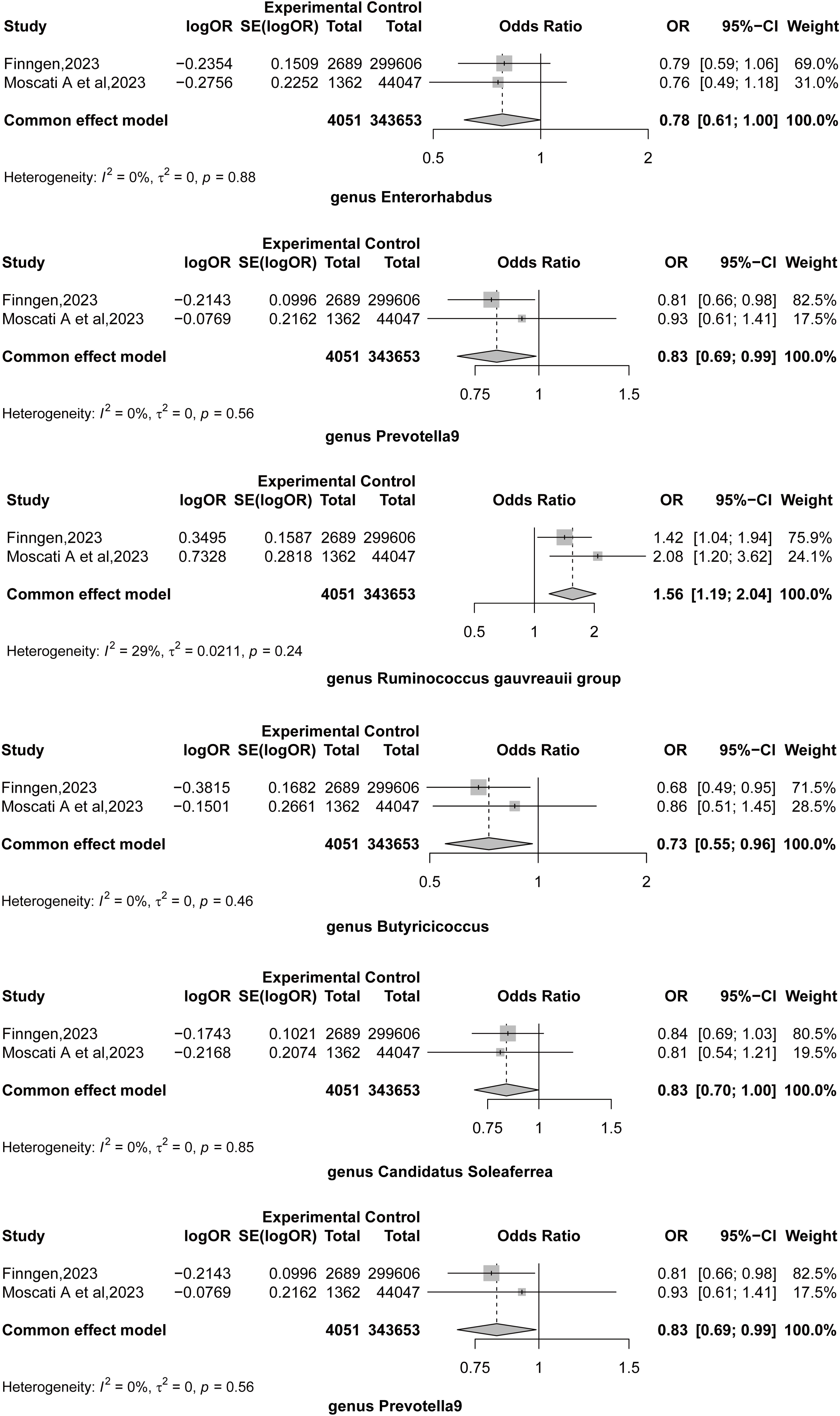
Meta results of gut microbiota and fibromyalgia mendelian randomization analysis.

**Table 1.** Inverse Mendelian randomization (MR) results of fibromyalgia and significant gut microbiota after meta-analysis.

| **Exposure** | **Outcome** | **Method** | **Nsnp** | **pval** |
| --- | --- | --- | --- | --- |
| Fibromyalgia | genus *Enterorhabdus* | Inverse variance weighted | 20 | 0.347 |
| Fibromyalgia | genus *Parabacteroides* | Inverse variance weighted | 20 | 0.601 |
| Fibromyalgia | genus *Ruminococcus gauvreauii* group | Inverse variance weighted | 20 | 0.219 |
| Fibromyalgia | genus *Butyricicoccus* | Inverse variance weighted | 20 | 0.202 |
| Fibromyalgia | genus *Candidatus Soleaferrea* | Inverse variance weighted | 20 | **0.004** |
| Fibromyalgia | genus *Prevotella 9* | Inverse variance weighted | 20 | 0.233 |

Nsnp: number of single nucleotide polymorphisms.

### 3.2. Causal relationship between 91 inflammatory proteins and fibromyalgia

In the preliminary analysis cohort, six inflammatory proteins showed statistically significant results (**Table S3**), including Natural killer cell receptor 2B4 (NKCR-2B4/CD244; OR = 1.18, 95% CI: 1.02-1.38, p = 0.031), CUB domain-containing protein 1 (OR = 0.78, 95% CI: 0.61-0.99, p = 0.037), S100-A12 (OR = 0.78, 95% CI: 0.61-0.99, p = 0.037), Interleukin-12 subunit beta (IL-12β; OR = 1.11, 95% CI: 1.02-1.21, p = 0.016), Monocyte chemoattractant protein 2 (MCP-2,/CCL8; OR = 0.90, 95% CI: 0.84-0.97, p = 0.008), and Tumor necrosis factor (TNF-ɑ; OR = 0.77, 95% CI: 0.61-0.96, p = 0.020). Significant immune proteins also passed sensitivity tests including Cochran’s Q test (p > 0.05), MR-Egger intercept test (p > 0.05), MR-PRESSO analysis (p > 0.05), and leave-one-out analysis (**Table S4, Figure S2**). MR analysis in the replication cohort showed similar trends for candidate immune proteins, but results were not significant due to large sample size differences.

We performed a meta-analysis of MR results from the preliminary and replication cohorts, ultimately identifying seven immune proteins influencing FM (**Figure 3**). Specifically, higher levels of C-X-C motif chemokine 5 (CXCL5; OR = 0.89, 95% CI: 0.80-1.00, p = 0.046), S100-A12 (OR = 0.76, 95% CI: 0.62-0.93, p = 0.007), Leukemia inhibitory factor receptor (LIFR; OR = 0.85, 95% CI: 0.73-0.98, p = 0.025), MCP-2/CCL8 (OR = 0.93, 95% CI: 0.87-0.99, p = 0.025), and TNF-α (OR = 0.79, 95% CI: 0.65-0.95, p = 0.013) reduced the risk of FM.

**Figure 3.**
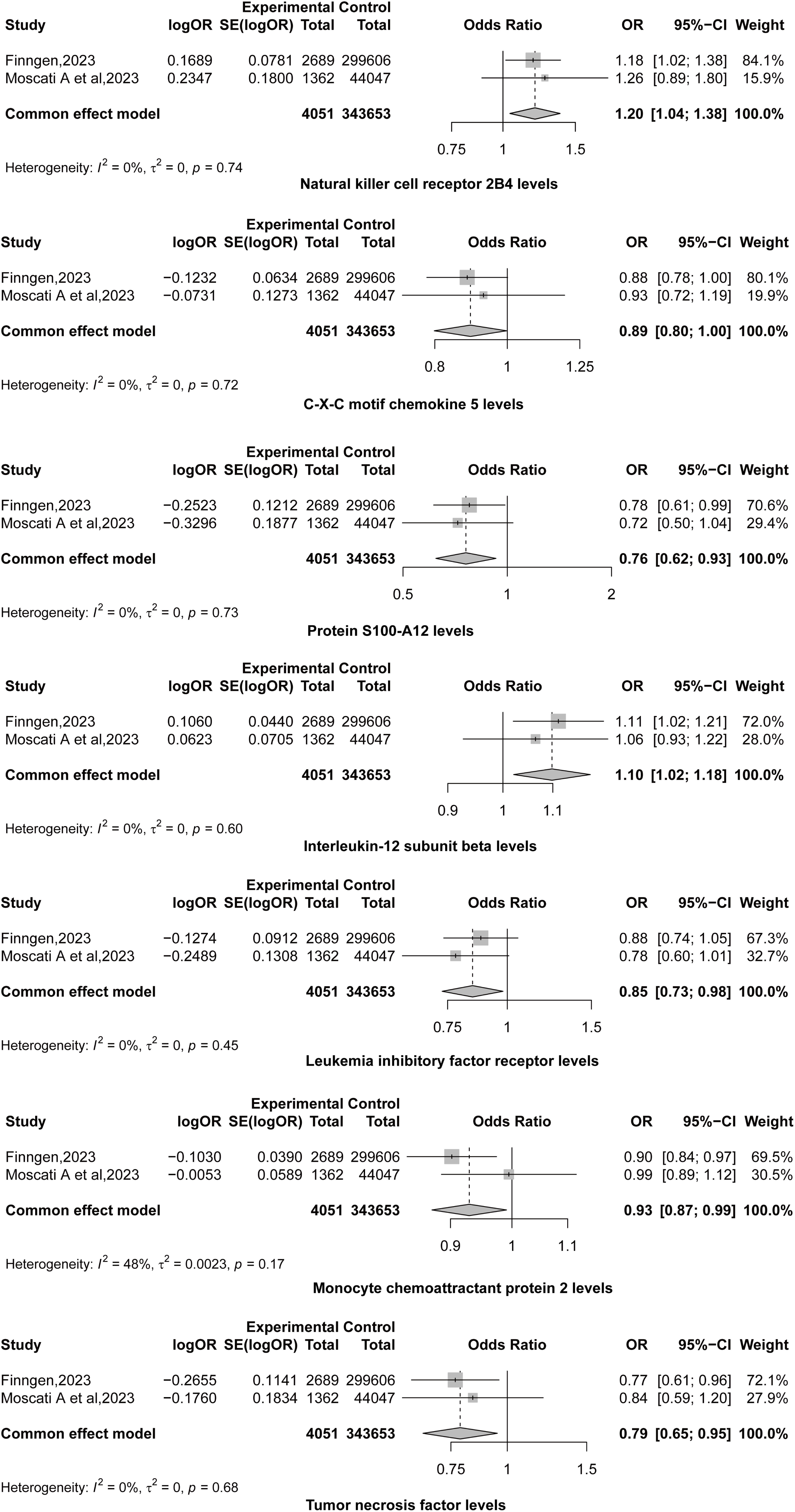
Meta results of inflammatory proteins and fibromyalgia mendelian randomization analysis.

However, higher levels of NKCR-2B4/CD244 (OR = 1.20, 95% CI: 1.04-1.38, p = 0.012) and IL-12β (OR = 1.10, 95% CI: 1.02-1.18, p = 0.012) increased susceptibility to FM. All significant results in the meta-analysis passed the reverse causality validation in subsequent analyses (**Table 2**).

**Table 2.** Inverse Mendelian randomization (MR) results of fibromyalgia and significant inflammatory protein after meta-analysis.

| **Exposure** | **Outcome** | **Method** | **Nsnp** | **pval** |
| --- | --- | --- | --- | --- |
| Fibromyalgia | Natural killer cell receptor 2B4 levels | Inverse variance weighted | 23 | 0.633 |
| Fibromyalgia | C-X-C motif chemokine 5 levels | Inverse variance weighted | 23 | 0.927 |
| Fibromyalgia | Protein S100-A12 levels | Inverse variance weighted | 23 | 0.520 |
| Fibromyalgia | Interleukin-12 subunit beta levels | Inverse variance weighted | 23 | 0.478 |
| Fibromyalgia | Leukemia inhibitory factor receptor levels | Inverse variance weighted | 23 | 0.141 |
| Fibromyalgia | Monocyte chemoattractant protein 2 levels | Inverse variance weighted | 23 | 0.910 |
| Fibromyalgia | Tumor necrosis factor (TNF-α) levels | Inverse variance weighted | 23 | 0.061 |

Nsnp: number of single nucleotide polymorphisms.

For the genes CXCL5, S100-A12, LIFR, CCL8, and TNF, the functional enrichment analyses using GO biological processes and KEGG pathways are shown in **Table S5**. According to results after False Discovery Rate (FDR), the GO analysis shows significant enrichment mainly in the “neutrophil chemotaxis.” KEGG pathway analysis indicates notable enrichment in “viral protein interaction with cytokine and cytokine receptor”. For the genes CD244 and IL-12β, the GO biological process and KEGG functional enrichment analyses are presented in **Table S6**. Post-FDR analysis reveals significant enrichment primarily in “positive regulation of interferon-gamma production” for GO biological processes, while KEGG analysis highlights significant enrichment in the “JAK-STAT signaling pathway.”

## 4. Discussion

### 4.1. Gut microbiota and fibromyalgia

Increasing evidence suggests that symbiotic gut bacteria play a significant role in FM (22–25). In our study, we identified *Enterorhabdus, Parabacteroides, Butyricicoccus,* and *Prevotella 9* as protective factors against FM, while the *R. gauvreauii* group was identified as a risk factor.

Research on *Enterorhabdus* is currently limited. A study investigating the antidepressant effects of Ziyan Green Tea on mildly stressed mice through fecal metabolomics revealed a significant negative correlation between *Enterorhabdus* and the pro-inflammatory cytokine IL-6 (41). This suggests that *Enterorhabdus* may help inhibit inflammatory responses, thereby acting as a protective factor. *Parabacteroides* has been shown to produce short-chain fatty acids (SCFAs), such as butyrate, which play critical roles in immune regulation (25, 42, 43). *Parabacteroides* may also help repair the intestinal barrier, reduce gut inflammation, and improve metabolic abnormalities like insulin resistance (44), highlighting its potential as a protective factor in FM. *Butyricicoccus* is one of the main producers of butyrate, a key immunomodulatory SCFA that inhibits dendritic cell differentiation and promotes the development of anti-inflammatory regulatory T cells, underscoring the potential role of *Butyricicoccus* in FM (23, 25, 45). Research on *Prevotella 9* in FM is scarce, but studies have suggested that *Prevotella* may act as a potential probiotic (46, 47). For example, in multiple sclerosis, treatment with *Prevotella histicola* increased the frequency and number of CD4FoxP3 regulatory T cells in the periphery and gut, while decreasing pro-inflammatory IFN-γ and IL17-producing CD4 T cells in the CNS, suggesting that *Prevotella histicola* suppresses disease progression by enhancing anti-inflammatory immune responses and inhibiting pro-inflammatory ones (47). Similarly, *Prevotella* may have protective properties through its anti-inflammatory activities.

*Ruminococcus*, on the other hand, may activate inflammatory signaling pathways through lipid dysregulation (especially PEA and glycerophosphocholine) and NLRP3 inflammasome activation (45). Importantly, *R. gauvreauii* may induce a systemic immune response by activating IL-6 and TNF-α signaling (48, 49). Moreover, *R. gauvreauii* may be associated with an increased risk for gastro-duodenal ulcers (50). Studies on *Ruminococcus* in depression have shown conflicting results. Zhao et al. found higher levels of *Ruminococcus* in major depressive disorder (MDD) patients, positively correlating with IL-1β, suggesting a pro-inflammatory role that may be important to the pathophysiology of FM (51). Conversely, other studies have shown higher relative abundance of *Ruminococcus* in healthy controls compared to MDD patients (52).

In 2007 (Maes et al., 2007), it was reported that the FM symptoms of chronic fatigue syndrome are associated with increased translocation of Gram-negative gut commensal bacteria and gut barrier dysfunction (22). Dysbiosis, particularly the imbalance between pathobionts (which promote inflammation and damage to epithelial cells and tight junctions) and symbionts (health-promoting microbiota, which engage in anti-inflammatory activities, support gut homeostasis and tight junctions, and produce SCFAs and vitamins), is a potential cause of gut barrier dysfunction and bacterial translocation, playing a crucial role in FM (53–56). Consequently, our results may suggest that FM is associated with a specific enterotype, namely with *R. gauvreauii* as a risk factor and *Enterorhabdus, Parabacteroides, Butyricicoccus,* and *Prevotella 9* as protective factors. These results deserve replication in new cohorts of patients with FM.

### 4.2. Cytokines/chemokines in fibromyalgia

In this study, five inflammatory proteins—CXCL5, S100-A12, LIFR, MCP-2/CCL8, and TNF-α—were identified as protective factors against FM. Conversely, NKCR-2B4/CD244 and IL-12β were identified as risk factors.

Functional enrichment analysis of CXCL5, S100-A12, LIFR, CCL8, and TNF genes revealed significant enrichment in the GO biological process “Cellular response to chemical stimulus,” with other genes also enriched in “Neutrophil chemotaxis,” “Leukocyte migration,” and “Cytokine-mediated signaling pathway”. KEGG pathway analysis indicated that CXCL5, CCL8, LIFR, and TNF genes were enriched in pathways such as “Viral protein interaction with cytokine and cytokine receptor,” “Cytokine-cytokine receptor interaction,” and “Chemokine signaling pathway”. For CD244 and IL-12β, GO biological process analysis showed enrichment in “Positive regulation of interferon-gamma production”; CD244 was also enriched in “Positive regulation of natural killer cell activation” and “Positive regulation of natural killer cell mediated immunity”. KEGG analysis revealed that CD244 was enriched in pathways like “JAK-STAT signaling pathway,” “Inflammatory bowel disease,” and “Th1 and Th2 cell differentiation”.

CXCL5, a CXCR2 ligand chemokine, is involved in recruiting leukocytes to tissues, contributing to neuroinflammation, blood-brain barrier breakdown, and neutrophil infiltration into the brain (57). Previous studies have shown that regulating the IL-6 pathway can alleviate pain-like behaviors in FM by reducing pro-inflammatory cytokines such as TNF-α and CXCL5. These observations may seem contradictory to our findings but reflect the complex interplay between immune activation and imbalance in FM, where both immune-inflammatory response system (IRS) and compensatory immune-regulatory response system (CIRS) are activated, leading to imbalances in regulatory and pro-inflammatory cytokines (4).

S100A12, a member of the S100 calcium-binding protein family, is mainly secreted by neutrophils, macrophages, and smooth muscle cells. Under inflammatory conditions, S100A12 binds to receptor for advanced glycation end-products (RAGE), signaling through JNK, ERK, and p38 to activate transcription factors AP-1 and nuclear factor (NF)-κB, leading to the release of pro-inflammatory cytokines (58). Elevated S100A12 levels have been observed in the saliva of FM patients (59). Previous studies also identified S100A12 as a diagnostic biomarker for rheumatoid arthritis and depression (60).

In a 2002 study by Maes et al, it was found that patients with depressive and anxiety symptoms during the early postpartum period exhibited reduced serum LIFR levels, which may contribute to immune activation (61). Another study indicated that long-term treatment with atypical antipsychotics could increase the anti-inflammatory capacity of serum in schizophrenia patients by elevating serum LIFR levels (62).

MCP-2/CCL8, a member of the CC chemokine subfamily, has been reported as an agonist for C-C chemokine receptor types 2 and 4. It plays a crucial role in regulating leukocyte chemotaxis and inflammatory diseases (57). A study by Bäckryd et al. found high levels of the chemokine CCL8 in the cerebrospinal fluid of patients with peripheral neuropathic pain compared to healthy controls (63). In a mouse study, MCP-2/CCL8 levels were increased in oxidative stress-induced allergic inflammation, and anti-inflammatory treatment was shown to reduce MCP-2/CCL8 levels (64). Another study revealed that MCP-2/CCL8 levels were higher in MDD patients with suicidal ideation (SI) compared to those without SI and healthy controls (65). MCP-2/CCL8 may contribute to the neurobiology of suicidality in depressed elderly individuals by enhancing neuroinflammatory responses, oxidative stress, apoptotic pathways, impairing neurogenesis, and disrupting serotonergic neurotransmission (65).

TNF-α is one of the most critical cytokines released by macrophages and microglia and is active in neuropathic pain pathways. It modulates pain signaling by binding to TNFR1 (TNF receptor 1) and plays a pivotal role in inflammation at pain sites (66). Research on TNF levels in FM patients has produced mixed results, with some studies reporting lower TNF levels (8, 67, 68) and others finding elevated TNF levels (69, 70).

CD244, also known as natural killer cell receptor 2B4, is a transmembrane protein belonging to the immunoglobulin superfamily and plays a vital role in immune regulation. KEGG analysis revealed that CD244 is primarily enriched in the JAK-STAT signaling pathway. Previous studies have shown that this pathway plays a significant role in FM pathogenesis, leading to chemokine overexpression and glial cell activation (66).

IL-12β is a pro-inflammatory cytokine and a component of IL-12, involved in activating Th1 immune responses. GO biological process analysis revealed that CD244, along with IL-12β, enhances inflammatory responses through the positive regulation of interferon-gamma production, which may be a risk factor for FM. Previous studies have found elevated IL-12 in patients with FM syndrome (66, 71). Recently, it has been shown that IL-12 also induces inflammatory hypernociception (72). A study demonstrated mechanical hypernociception in rats injected with IL-12, mediated by endothelin-1 and endothelin-B receptor interaction. This finding is significant as pain at the IL-12 injection site has been reported in human treatment (73). IL-12 has also been identified as an essential pro-inflammatory factor in other diseases, such as COVID-19, major depression, psoriasis, and systemic lupus erythematosus (74–78).

To summarize, the immunological characteristics observed in the present investigation of FM cannot be solely attributed to a simple “inflammation.” The development of FM may be assigned to an intricate imbalance between the IRS and the CIRS. A recent meta-analysis has substantiated that FM is distinguished by alterations and disparities in immunoregulatory and pro-inflammatory cytokines, resulting in peripheral immunological activation (4). Based on the analysis of enrichment and annotation, the present study indicates that FM may be associated with elevated production of IFN-γ and activation of natural killer cells, as well as reduced chemotaxis and diminished defense against viruses. Indeed, CXCL5, CCL8, and TNF participate in the hsa04061 KEGG pathway (KEGG PATHWAY: hsa04061 (genome.jp)), specifically referred to as “viral protein interaction with cytokine and cytokine receptor”. DNA viruses could undermine the host immune system network in this pathway. A recent study by Maes et al. (2024) demonstrated a significant association between the replication of human herpes type 6 (HHV-6) and the manifestation of FM symptoms in individuals with Long COVID. In addition, reactivation of HHV-6 can potentially impact the integrity of the gut tight junctions, resulting in immune-inflammatory reactions (79). Another report indicated that individuals suffering from FM may exhibit HHV-6 infection (80). Consequently, the replication of HHV-6 may play a role in the immune-inflammatory mechanisms associated with FM by activation the immune-inflammatory response system (81), impairing chemotaxis and inducing antiviral responses via elevated production of IFN-γ (82). Consequently, the elevation of IFN-γ production has the potential to elicit peripheral pain responses via interactions between neurons and microglia within the spinal cord (82, 83). The immune response in FM may be exacerbated by the heightened pro-inflammatory effects of *R. gauvreauii*, alongside a diminished presence of gut bacteria that typically provide a protective barrier against inflammation and the phenomenon of leaky gut. This includes notable genera such as *Enterorhabdus, Parabacteroides, Butyricicoccus,* and *Prevotella 9*.

### 4.3. Limitations

When interpreting the results of this study, certain limitations must be acknowledged. This study was conducted on a European population, which may introduce ethnic variability in the findings. Further validation is required through cellular, animal, and clinical studies.

## 5. Conclusion

This study presents a comprehensive bidirectional MR analysis and meta-analysis investigating the causal relationships between gut microbiota, inflammatory proteins, and FM. Our findings identify *R. gauvreauii* as a potential risk factor for FM, while *Enterorhabdus*, *Parabacteroides*, *Butyricicoccus*, and *Prevotella 9* appear to act as protective factors. Additionally, five inflammatory proteins—CXCL5, S100-A12, LIFR, MCP-2/CCL8, and TNF-α —were associated with protective roles, whereas NKCR-2B4/CD244 and IL-12β were identified as risk factors. The study provides new insights into the complex interactions between gut microbiota, immune responses, and FM pathophysiology. The dysregulation of immune-inflammatory and compensatory immune-regulatory systems appears central to FM development, with potential links to viral pathways such as HHV-6 reactivation. In conclusion, this study underscores the importance of gut microbiota and immune modulation in FM, offering promising avenues for future therapeutic interventions.

### Ethical statement

Our analyses were based on publicly available data that had been approved by relevant review boards. Therefore, no additional ethical approval is required.

### Funding

There was no specific funding for this specific study.

## Acknowledgment

The research community is thanked for making summary statistics from genome-wide association studies publicly available.

## Author’s contributions

All the contributing authors have participated in the preparation of the manuscript.

## Data Access Statement

The datasets used and/or analyzed during the current study are presented in the manuscript. Summary statistics for GWAS are publicly available.

