## Supplementary Figure 1-2 for "A multi-omics bidirectional mendelian randomization study and meta-analysis on the causal relationship between gut microbiota, inflammatory proteins, and fibromyalgia"

**Figure S1 Leave-one-out analysis of gut microbiota with significant MR analysis results and fibromyalgia (preliminary analysis cohort).**

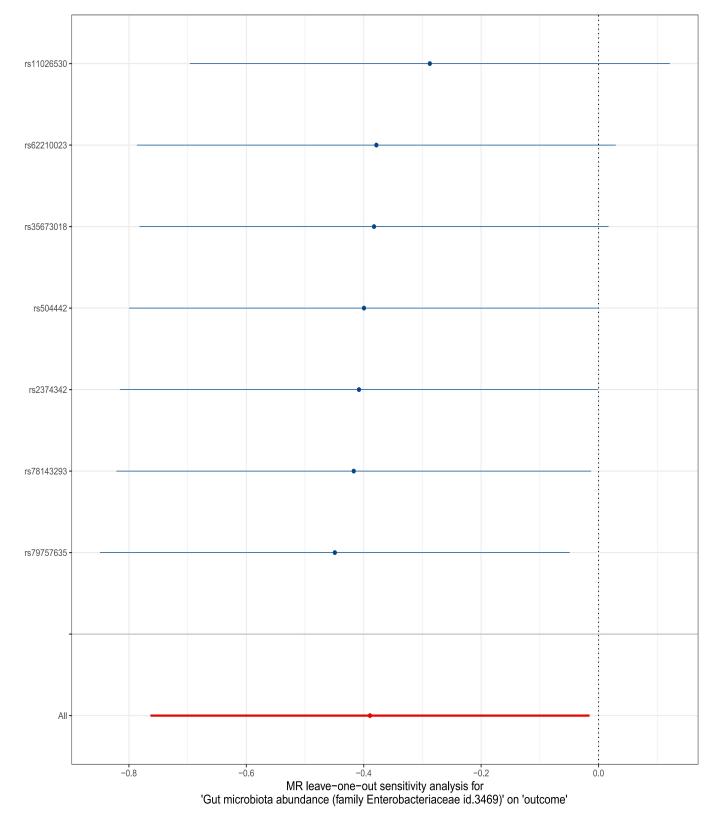

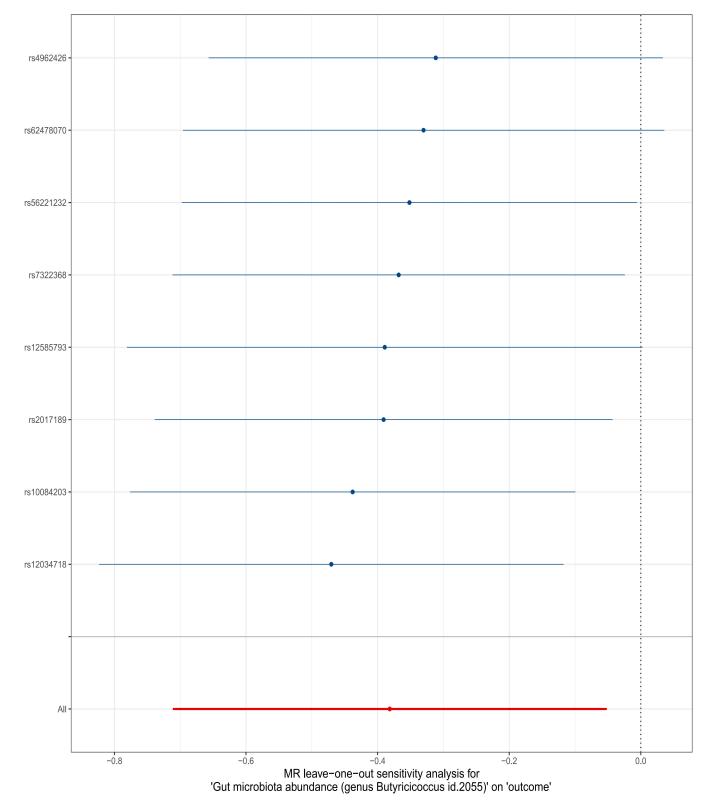

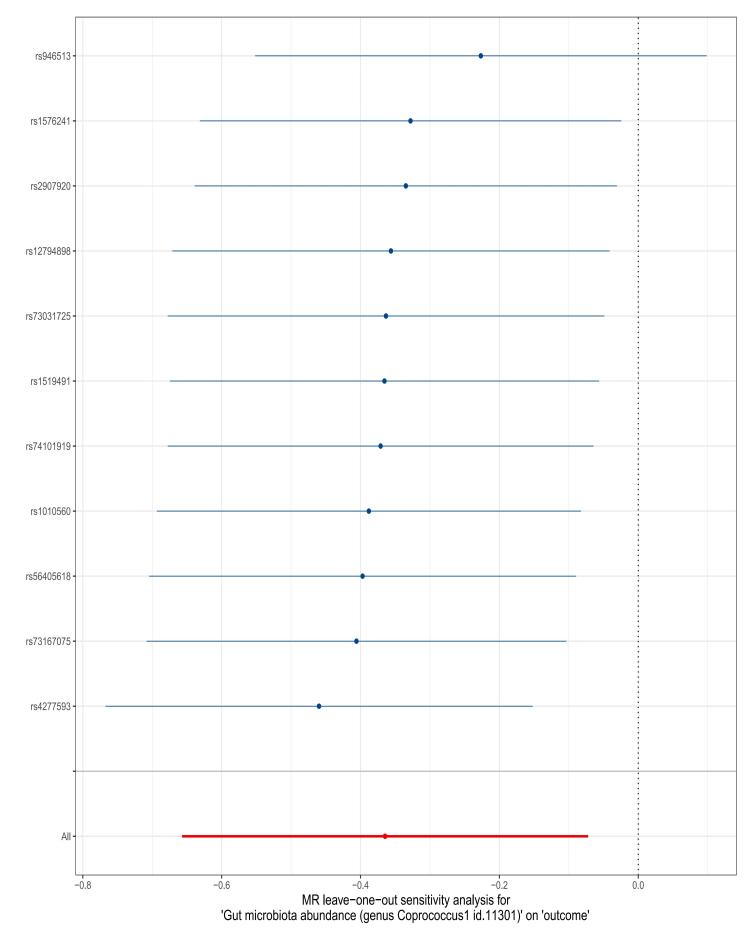

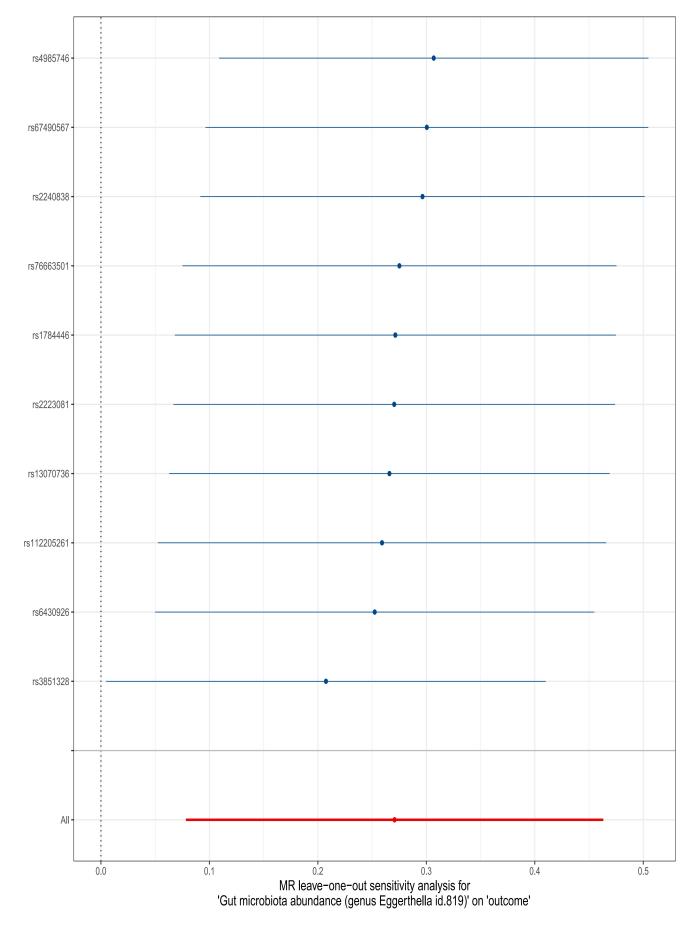

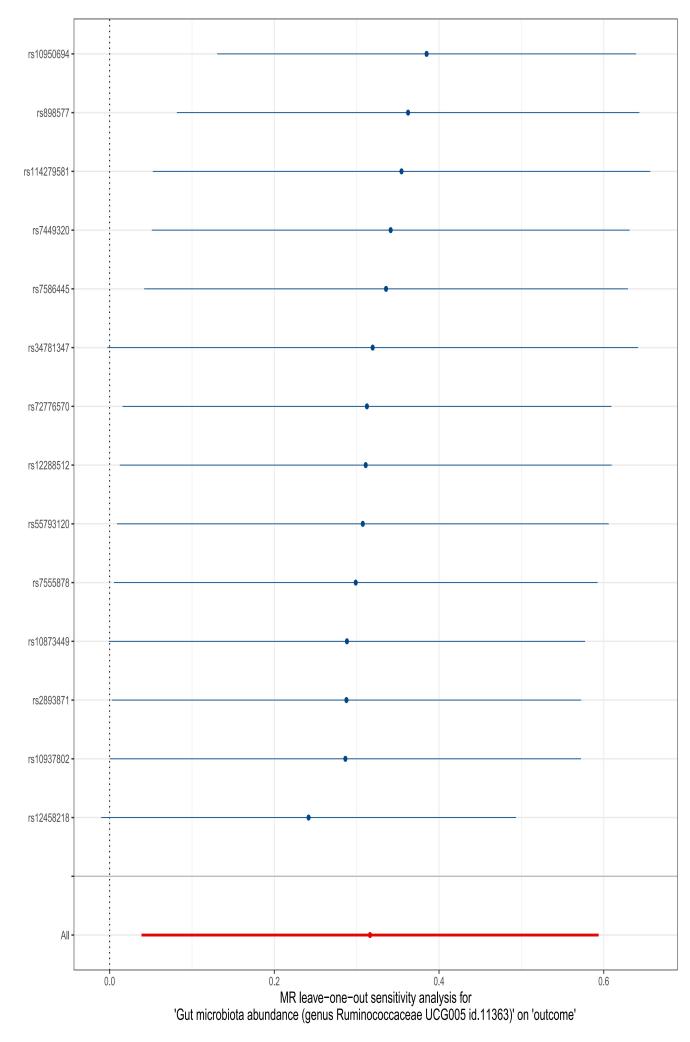

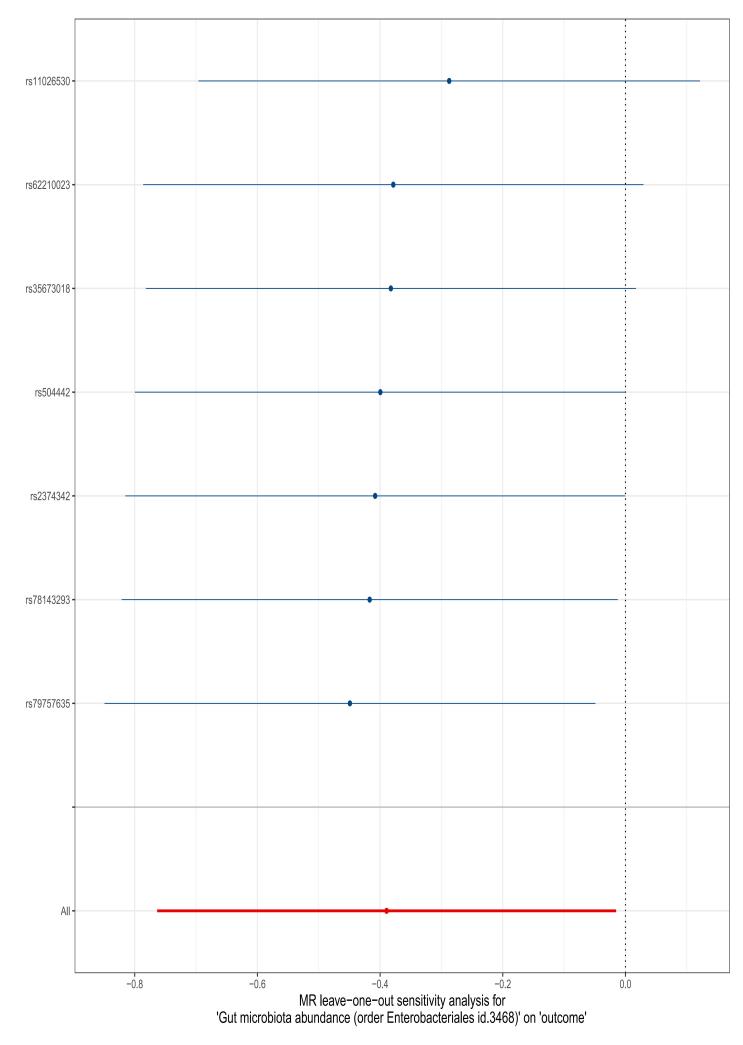

**Figure S2 Leave-one-out analysis of inflammatory protein with significant MR analysis results and fibromyalgia (preliminary analysis cohort).**

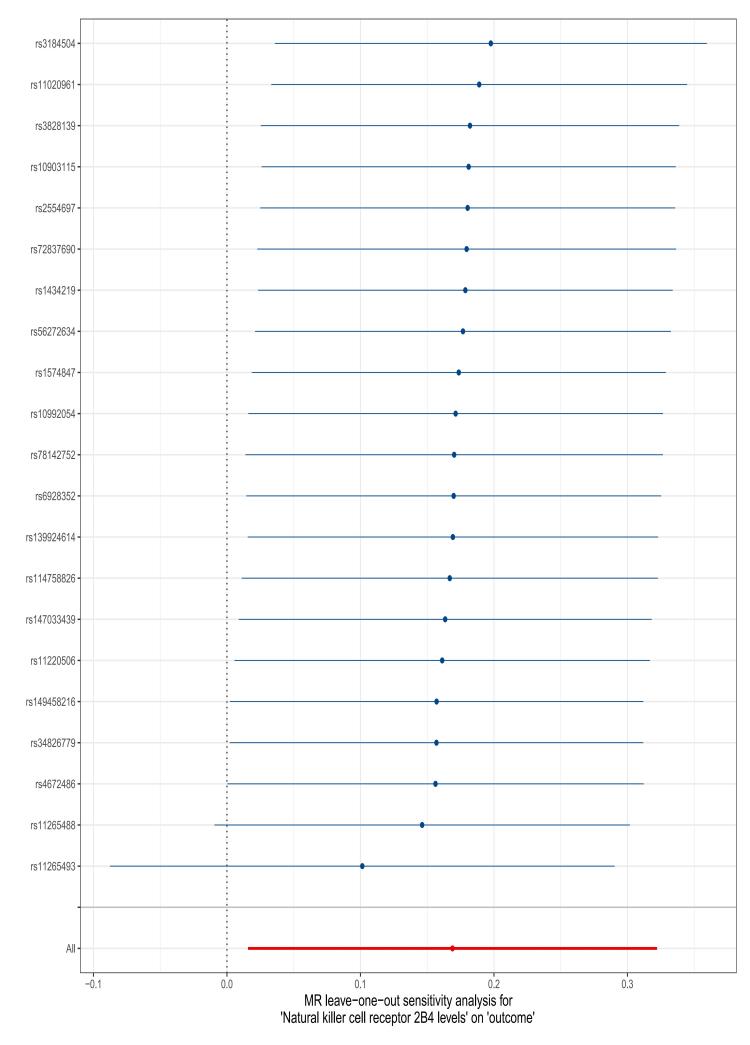

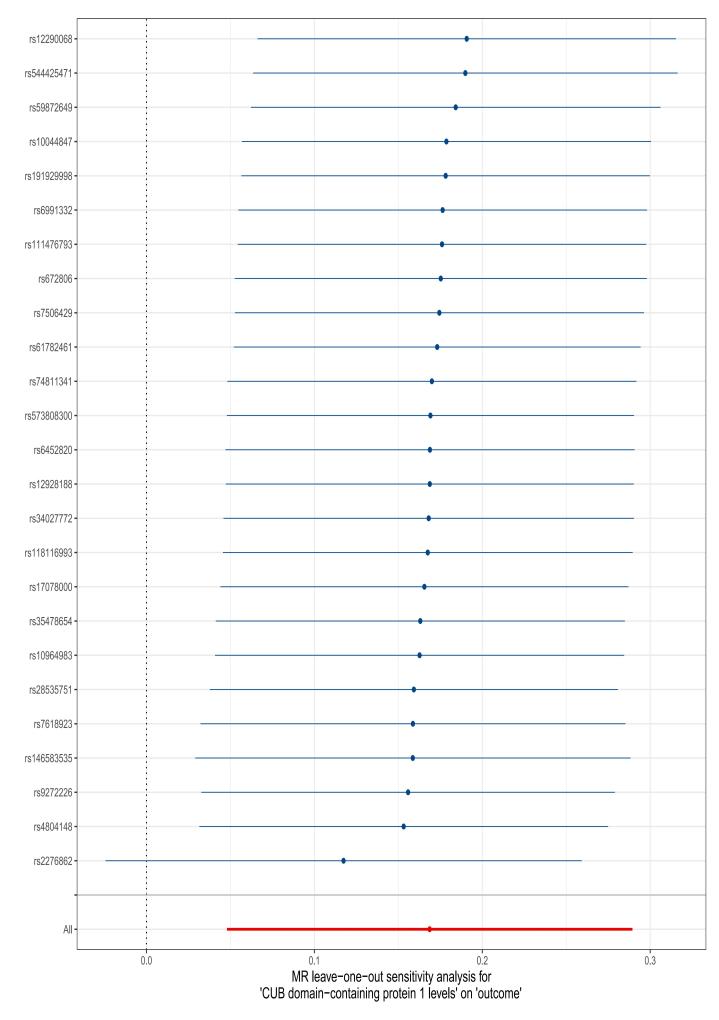

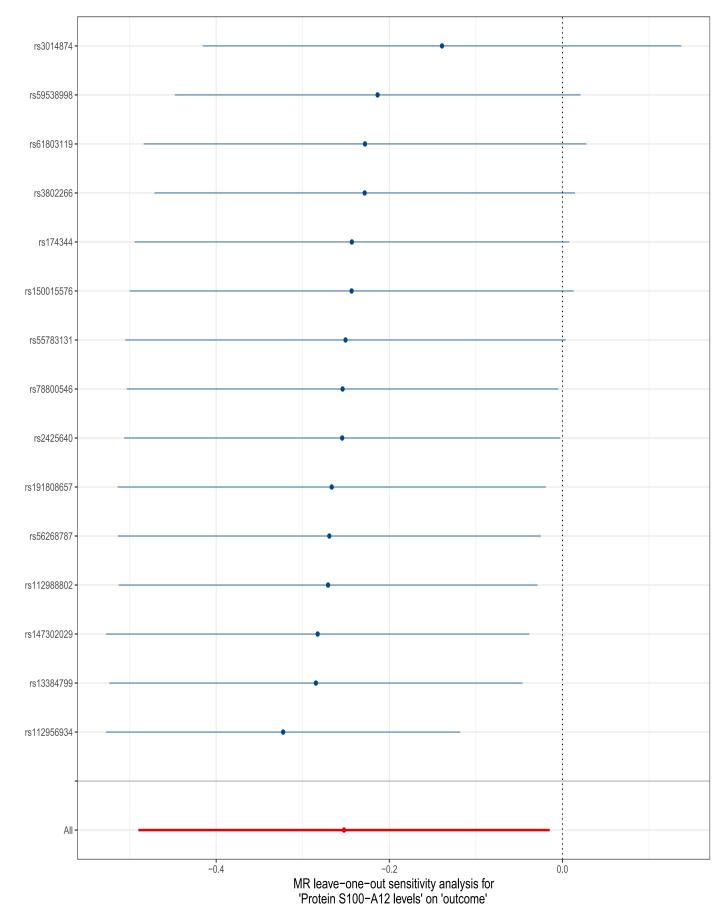

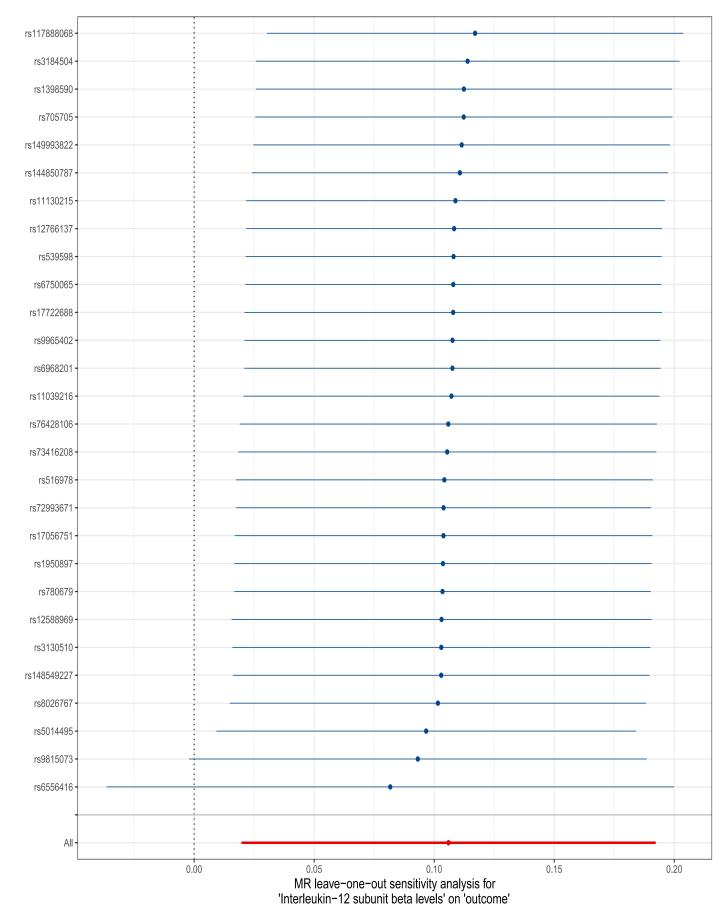

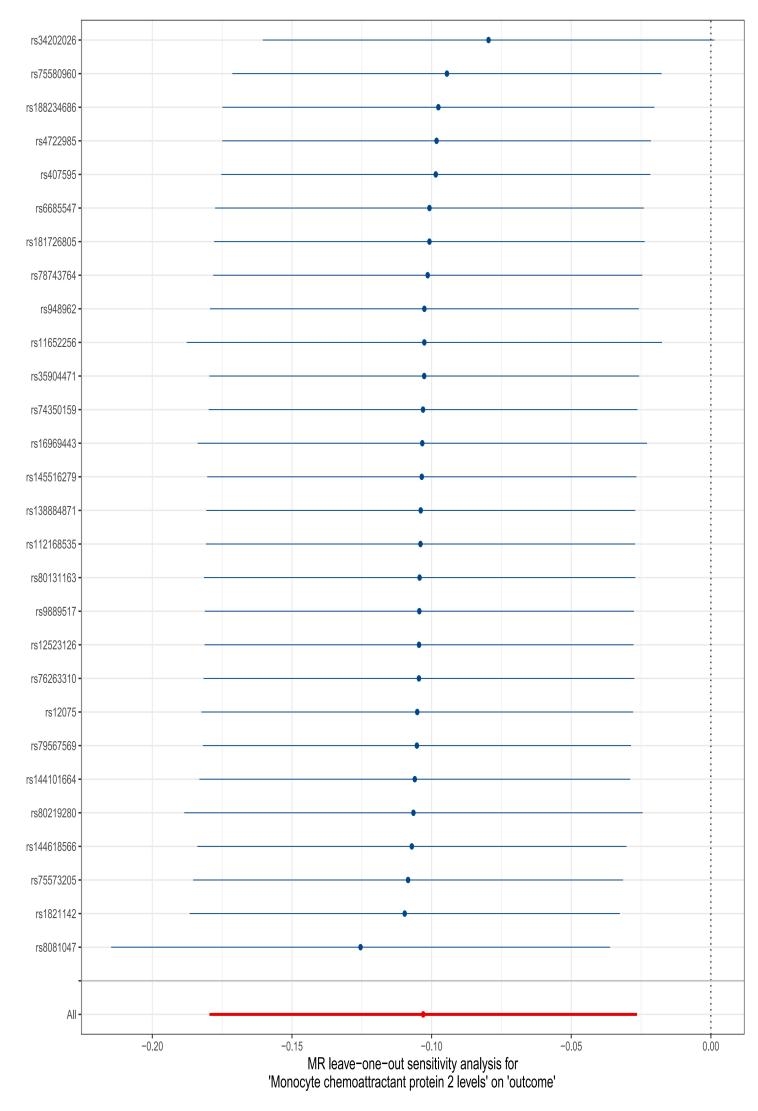

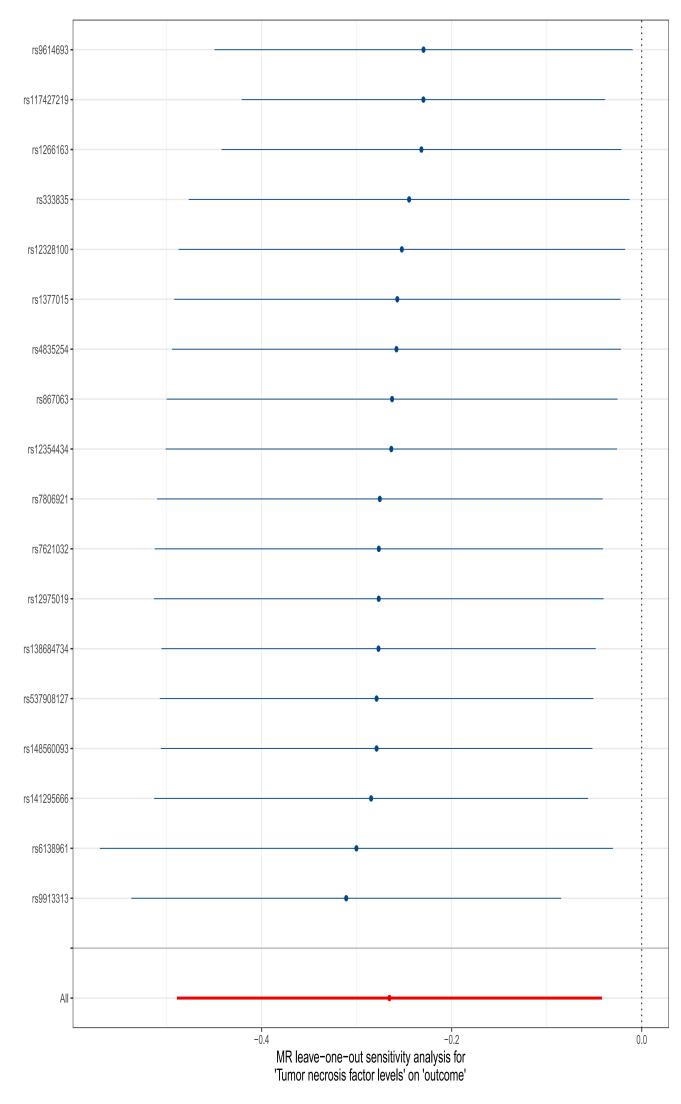
